# Evaluating algorithmic fairness in the presence of clinical guidelines: the case of atherosclerotic cardiovascular disease risk estimation

**DOI:** 10.1101/2021.11.08.21266076

**Authors:** Agata Foryciarz, Stephen R. Pfohl, Birju Patel, Nigam H. Shah

## Abstract

The American College of Cardiology and the American Heart Association guidelines on primary prevention of atherosclerotic cardiovascular disease (ASCVD) recommend using 10-year ASCVD risk estimation models to initiate statin treatment. For guideline-concordant decision making, risk estimates need to be calibrated. However, existing models are often miscalibrated for race, ethnicity, and sex based subgroups. This study evaluates two algorithmic fairness approaches to adjust the risk estimators (group recalibration and equalized odds) for their compatibility with the assumptions underpinning the guidelines’ decision rules. Using an updated Pooled Cohorts dataset, we derive unconstrained, group-recalibrated, and equalized odds-constrained versions of the 10-year ASCVD risk estimators, and compare their calibration at guideline-concordant decision thresholds. We find that, compared to the unconstrained model, group-recalibration improves calibration at one of the relevant thresholds for each group, but exacerbates differences in false positive and false negative rates between groups. An equalized odds constraint, meant to equalize error rates across groups, does so by miscalibrating the model overall and at relevant decision thresholds. Hence, because of induced miscalibration, decisions guided by risk estimators learned with an equalized odds fairness constraint are not concordant with existing guidelines. Conversely, recalibrating the model separately for each group can increase guideline compatibility, while increasing inter-group differences in error rates. As such, comparisons of error rates across groups can be misleading when guidelines recommend treating at fixed decision thresholds. The illustrated tradeoffs between satisfying a fairness criterion and retaining guideline compatibility underscore the need to evaluate models in the context of downstream interventions.

**SUMMARY:** 

**What is already known?:**

- Algorithmic fairness methods can be used to quantify and correct for differences in specific model performance metrics across groups, but the choice of an appropriate fairness metric is difficult.
- The Pooled Cohort Equations (PCEs), 10-year ASCVD risk prediction models used to guide statin treatment decisions in the United States, exhibit differences in calibration and discrimination across demographic groups, which can lead to inappropriate or misinformed treatment decisions for some groups
- Two theoretically incompatible fairness adjustments have been separately proposed for re-deriving the PCEs

**What does this paper add?:**

- Proposes a measure of local calibration of the PCEs at therapeutic thresholds as a method for probing guideline compatibility
- Quantifies the effect of two proposed fairness methods for re-deriving the PCEs in terms of their impact on local calibration
- Illustrates general principles that can be used to conduct contextually-relevant fairness evaluations of models used in clinical settings in the presence of clinical guidelines

## INTRODUCTION

While risk stratification models are central to personalizing care, their use can worsen health inequities [1]. In an effort to mitigate harms, several recent works propose *algorithmic group fairness* - mathematical criteria which require that certain statistical properties of a model’s predictions not differ across groups [2,3]. However, identifying which statistical properties are most relevant to fairness in a given context is non-trivial. Hence, before applying fairness criteria for evaluation or model adjustment, it is crucial to examine how the model’s predictions will inform treatment decisions - and what effect those decisions will have on patients’ health. Here, we consider the 2019 guidelines of the American College of Cardiology and the American Heart Association (ACC/AHA) on primary prevention of atherosclerotic cardiovascular disease (ASCVD) [4], which codify the use of 10-year ASCVD risk predictions to inform a clinician-patient shared decision making on initiating statin therapy. These guidelines recommend that individuals estimated to be at intermediate risk (>7.5-20%) be considered for initiation for moderate- to high-intensity statin therapy, and that those at high risk (>20%) be considered for high-intensity statin therapy. Individuals at borderline risk (>5-7.5%) may be considered for therapy under some circumstances [4,5].

These *therapeutic thresholds* were established based on randomized control trials, and correspond to risk levels where expected overall benefits derived from LDL cholesterol reduction outweigh risks of side effects (Supplement C) [4,6]. In general, such thresholds can be identified using decision analysis methods [7] (Figure 1A). The models accompanying the guidelines (Pooled Cohort Equations, PCEs [6,8,9]), developed for Black women, white women, Black men and white men, differ in both calibration and discrimination across groups [10,11]. The resultant systematic bias in risk misestimation in these subgroups can lead to inappropriate or misinformed treatment decisions. Since then, several works derived updated equations [11–14], some explicitly incorporating fairness adjustments [13,14].

**Figure 1.**
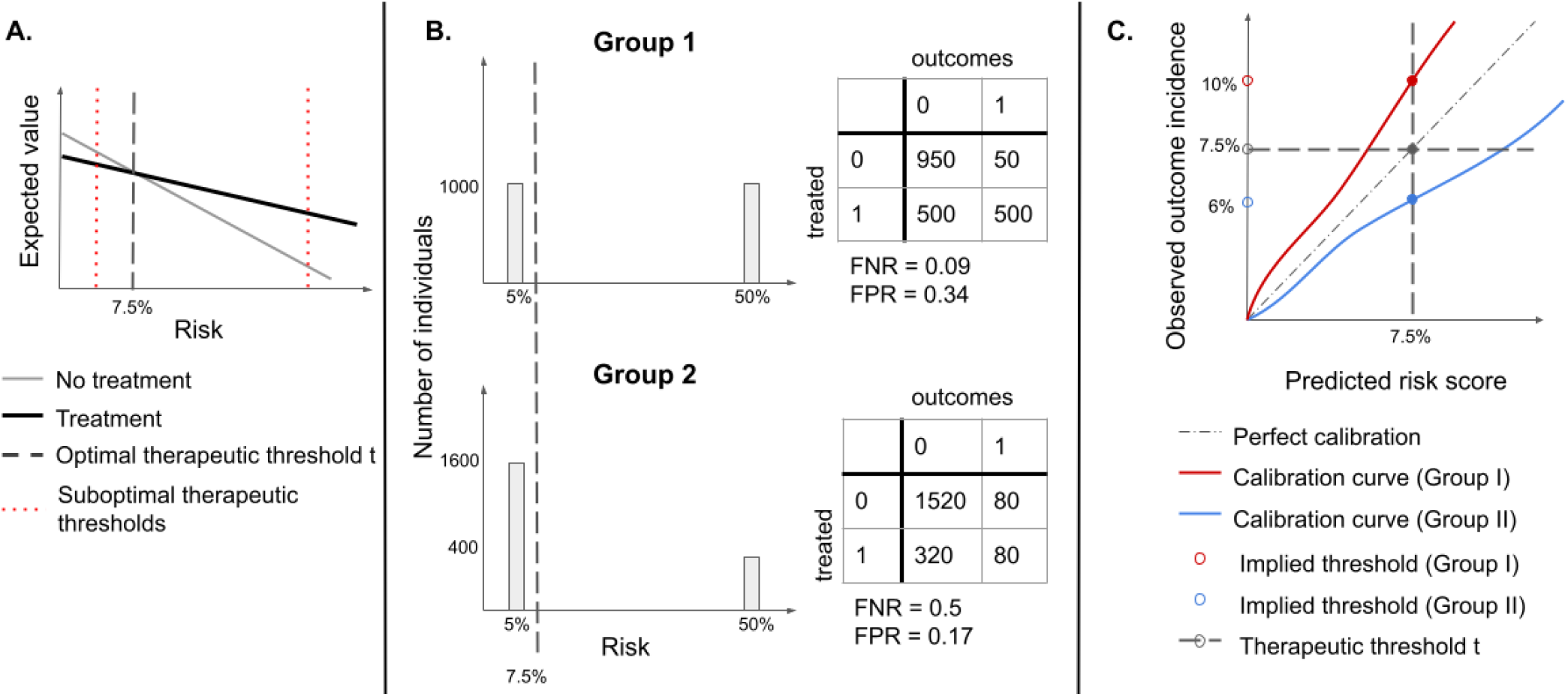
A. Identifying an optimal therapeutic threshold. An individual with risk *r* should be treated if the expected value of treatment exceeds that of non-treatment. As risk increases, the benefits of treatment become more significant, and assigning treatment becomes more optimal than withholding it. The optimal therapeutic threshold t is the value of risk at which treatment and non-treatment have the same expected value (the indifference point) - for individuals with *r*>t, treatment is expected to be more beneficial than non-treatment. Setting a non-optimal therapeutic threshold could lead to suboptimal treatment decisions for some individuals (treating some individuals for whom non-treatment has a higher expected value, or not treating individuals for whom treatment has a higher expected value). **Figure 1B. Illustration of the problem of infra-marginality**. Assume that there are two types of easily distinguishable individuals: with 5% and 50% chance of developing a disease respectively, and there are two groups composed of both types of individuals, but one has a higher proportion of lower-risk individuals. If the same therapeutic threshold is applied to both groups, False Positive Rates (FPR) and False Negative Rates (FNR) will not be equal, even though we would be making optimal treatment decisions for each patient, in both populations. **Figure 1C. Under miscalibration, implied thresholds differ from therapeutic thresholds**. If risk scores are miscalibrated, taking action at the threshold of 7.5% corresponds to different observed outcome rates in the two groups. For Group I, a risk score of 7.5% corresponds to an observed outcome incidence of 10%, while for Group II it corresponds to 6%, therefore, individuals in Group II would be treated at a lower risk than individuals in Group I.

If therapeutic thresholds recommended by guidelines reflect a balance of relevant harms and benefits for all subgroups [15,16], therapeutic decisions could be unfair if thresholds used for different groups differ, as they would lead to suboptimal treatment decisions for some groups (Figure 1A). As such, subgroup calibration at optimal therapeutic thresholds is an important fairness criterion for 10-year ASCVD risk estimation models [14], since under miscalibration (systematic over- or under-estimation of risk), treatment thresholds implicitly change (Figure 1C) from treatment thresholds to *implied thresholds* [17,18].

An alternative fairness criterion, known as *equalized odds* [3], which has previously been used to evaluate several clinical predictive models [13,19,20], requires equality in false positive and false negative error rates (FPR, FNR) across groups. One work proposed to explicitly incorporate equalized odds constraints into the training objective to learn ASCVD risk estimators with minimal inter-group differences in FPR and FNR [13].

In the context of ASCVD risk estimation, the equalized odds criterion lacks a clear motivation and can thus yield misleading results. FPR and FNR are sensitive to the distribution of risk and are expected to differ across groups when the incidence of outcomes differs (Figure 1B) [18,21,22]. Furthermore, approaches to build equalized odds-satisfying models either explicitly adjust group-specific decision thresholds, introduce differential miscalibration, or reduce model fit for each group [3] - which may lead to suboptimal decisions (Figure 1A, C). Equalized odds-satisfying models may therefore be less appropriate than calibrated estimators for use with the ACC/AHA guidelines [17].

We aim to evaluate the tension between calibration, equalized odds, and guideline-concordant decision making. To do so, we propose a measure of local calibration at guideline-concordant therapeutic thresholds as a method for probing guideline compatibility and apply it to unconstrained, group-recalibrated, and equalized odds-constrained versions of the 10-year ASCVD risk prediction models learned from the updated Pooled Cohorts dataset [9,11], as well as the original [8] and revised PCEs [11]. We assess the proposed local calibration measure and error rates across groups for each model, and conclude with recommendations for identifying quantification and adjustment criteria for enabling fair model-guided decisions.

## METHODS

### Datasets

We use an updated Pooled Cohorts dataset [11], comprised of ARIC (Atherosclerosis Risk in Communities Study, 1987-2011), CARDIA (Coronary Artery Risk Development in Young Adults Study, 1983-2006), CHS (Cardiovascular Health Study, 1989-1999), FHS OS (Framingham Heart Study Offspring Cohort, 1971-2014), MESA (Multi-Ethnic Study of Atherosclerosis, 2000-2012) and JHS (Jackson Heart Study, 2000-2012). Following the original PCE inclusion criteria [9], we include individuals aged 40-79, excluding those with a history of myocardial infarction, stroke, coronary bypass surgery, angioplasty, congestive heart failure or atrial fibrillation, or receiving statins at the time of the initial exam. We include all individuals, regardless of racial category, and classify them as Black and non-Black, consistent with the use of PCEs in practice for non-Black patients of color.

We extract features included in the PCEs (total cholesterol, HDL cholesterol, treated and untreated systolic blood pressure, diabetes, smoking status, age, binary sex and race), and BMI, recorded at the initial examination. We also extract dates of observed ASCVD events (myocardial infarction, lethal or non-lethal stroke or lethal coronary heart disease), and of last recorded observation (follow up or death), to define binary labels for 10-year ASCVD outcome and censoring. Individuals whose last recorded observation happened before an ASCVD event and before year ten are considered censored. We remove records with extreme values of systolic blood pressure (outside 90-200 mmHg), total cholesterol and HDL cholesterol (outside 130-320 and 20-100 mg/dL, respectively) or missing covariates.

### Models

#### Unconstrained model

The original PCEs consist of four separate Cox proportional hazards models, stratified by sex and race, to account for differences in ASCVD incidence across the four groups (Black women, white women, Black men and white men) [8]. One revision of the PCEs, which reduced overfitting and improved calibration, replaced the Cox models with censoring-adjusted logistic regression models, stratified by sex, and included race as a variable in each model [11]. Our implementation of the unconstrained models consists of a single inverse probability of censoring (IPCW)-adjusted logistic regression model [23], and includes race and sex as binary variables. Censoring weights are obtained from four group-level Kaplan-Meier estimators applied to the training set. We include all features and their two-way interactions.

#### Group-recalibrated model

For recalibration, we logit-transform the predicted probabilities generated by the unconstrained model, and use IPCW-adjusted logistic regression to fit a calibration curve for each group. We then use the resulting group-recalibrated model to obtain a set of recalibrated predictions.

#### Equalized odds model

The equalized odds criterion requires that both the FPR and FNR be equal across groups at one or more thresholds [3]. We use an in-processing method for constructing equalized odds models [22,24], which provides a better calibration-equalized odds tradeoff than the post-processing approach [25]. We define the training objective by adding a regularizer to the unconstrained model’s objective (Supplement A), with the degree of regularization controlled by λ. The regularizer penalizes differences between FPR and FNR at specified decision thresholds (7.5%, 20%), across the four groups.

### Training procedures

Using random sampling stratified by group, outcome and presence of censoring, we divide our cohort into the training (80%), recalibration (10%) and test (10%) sets. Using the same procedure, we divide the training set into 10 equally-sized subsets and, for each subset, train a logistic regression model using stochastic gradient descent for up to 200 iterations of 128 minibatches, with learning rate of 10^−4^ on the remaining subsets. We terminate training if the cross-entropy loss does not improve on the held-out subset for 30 iterations. This procedure generates ten unconstrained models. To generate group-recalibrated models, we first generate predictions on the recalibration set, using the unconstrained models (Figure 2), and then use those train logistic regression models using BFGS optimization, implemented in Scikit-Learn [26], with up to 10^5^ iterations. To examine the impact of the equalized odds penalty, we repeat the unconstrained training procedure using the regularized training objective with four different settings of the parameter λ, distributed log-uniformly on the interval 0.1-1.0 (0.100, 0.215, 0.464, 1.000), and refer to the resulting models as EO1 through EO4. Pytorch version 1.5.0 [27] is used to define all models and training procedures.

**Figure 2.**
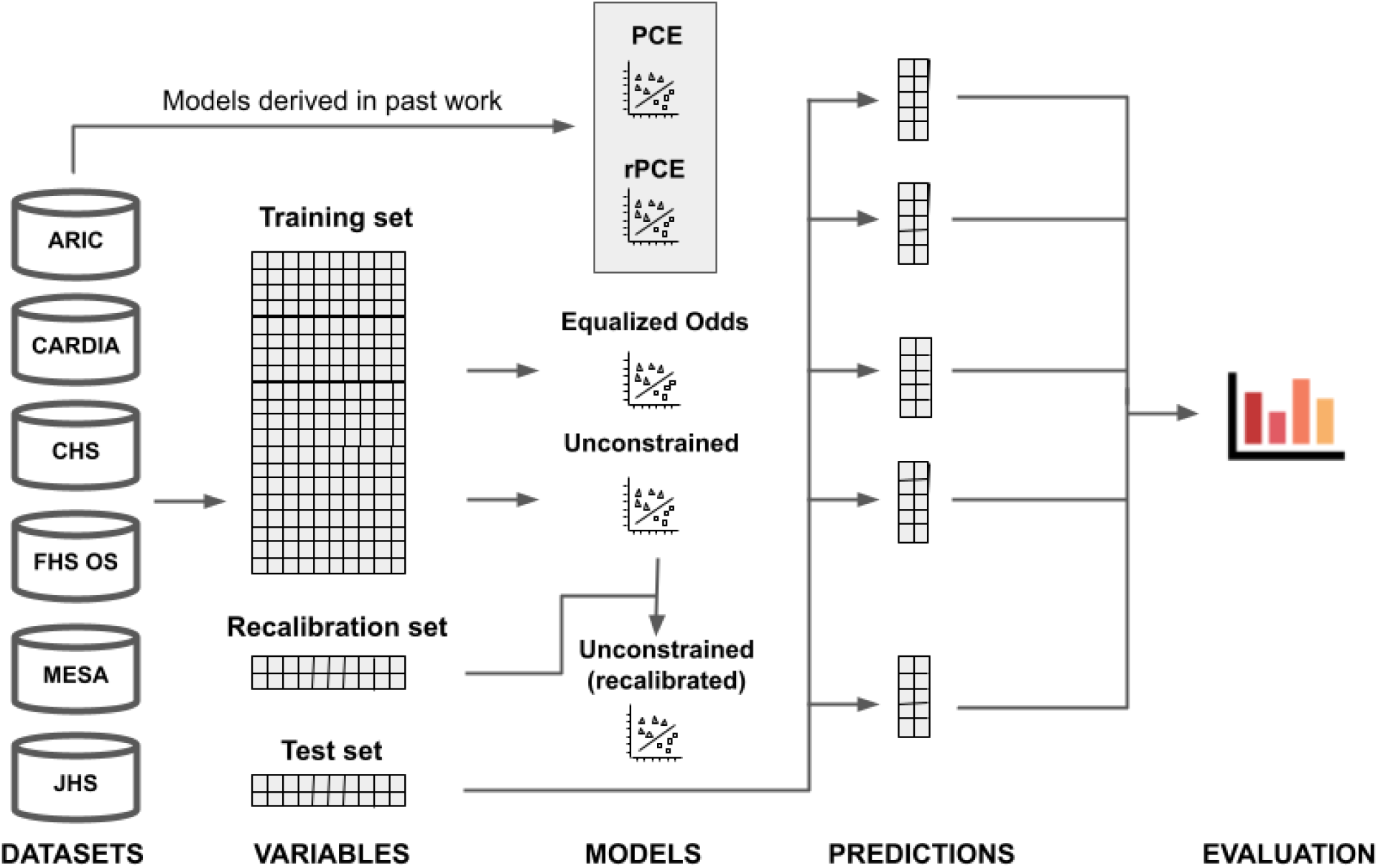
Visual abstract. Data from the 6 considered datasets: ARIC (Atherosclerosis Risk in Communities Study), CARDIA (Coronary Artery Risk Development in Young Adults Study), CHS (Cardiovascular Health Study), FHS OS (Framingham Heart Study Offspring Cohort), MESA (Multi-Ethnic Study of Atherosclerosis), and JHS (Jackson Heart Study), is extracted using the cohort definition used in the original PCEs, and divided into train (80%), validation (10%) and test (10%) sets. Equalized odds (EO) and unconstrained (UC) models are derived directly from the training set. The recalibrated model (rUC) is derived from the unconstrained model using a recalibration procedure, which uses the validation dataset (not seen during training). Finally, predictions on the test set are generated for all models - including the PCEs and the revised PCEs (rPCE), derived in past work - and evaluated.

### Evaluation

We introduce *threshold calibration error* (TCE), a measure of local calibration, defined as the difference between the therapeutic threshold (t_1_=7.5% or t_2_=20%) applied on the risk estimate and the *implied threshold* on the risk, measured by the calibration curve (Figure 1C). As in the recalibration procedure, we estimate implied thresholds *g*_a_(t_i_) at a fixed therapeutic threshold t_i_ by fitting a calibration curve *g*_a_ for each group *a* (Figure 1C). Then, for each threshold *i* we obtain TCE_ia_:

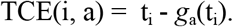

A negative TCE indicates risk underestimation, since the threshold applied to the risk score is lower than the observed incidence of the outcome at that predicted risk level. Similarly, a positive TCE indicates risk overestimation.

To understand the trade-off between TCE, FPR and FNR, we calculate inter-group standard deviation (IGSD) between the four group-specific values of the three metrics. For a threshold *i*, metric *M*, and *A* distinct groups, IGSD_*Mi*_ is defined as

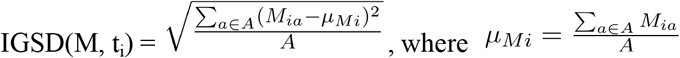

IGSD captures the degree of performance disparity between groups; high IGSD in FPR and FNR corresponds to an equalized odds disparity, and high IGSD in TCE corresponds to a treatment rule disparity. For each of the four subgroups, and overall population, we report calibration and discrimination metrics at both the aggregate (Absolute Calibration Error, ACE [22] and AUROC) and the threshold level (TCE, FPR, FNR) at t_1_ and t_2_, for the unconstrained model (UC), the group-recalibrated model (rUC), and the best-performing equalized odds model (EO), as well as the original PCEs [9] (PCE), and revised PCEs [11] (rPCE). We draw 1,000 bootstrap samples from the test set, stratified by group and outcomes, to derive point estimates and 95% confidence intervals (CI) for each metric. The CIs are defined as the 2.5% and 97.5% percentiles of the distribution obtained via pooling over both the bootstrap samples and the ten model replicates derived from the training procedure. We also report IGSD between the four group-specific median values in TCE, FPR and FNR at both thresholds. All metrics are computed over the uncensored population and adjusted for censoring using IPCW.

## RESULTS

We describe the study population and present performance of the models. We report the TCE, FPR and FNR in Figure 3, and IGSD of those three metrics in Figure 4. We present results for EO3 in Figure 3, as it was the only equalized odds model that achieved a reduction of IGSD(FPR) while keeping a low IGSD(FNR) at both thresholds. Results for the remaining equalized odds models are included in Supplement B.

**Figure 3.**
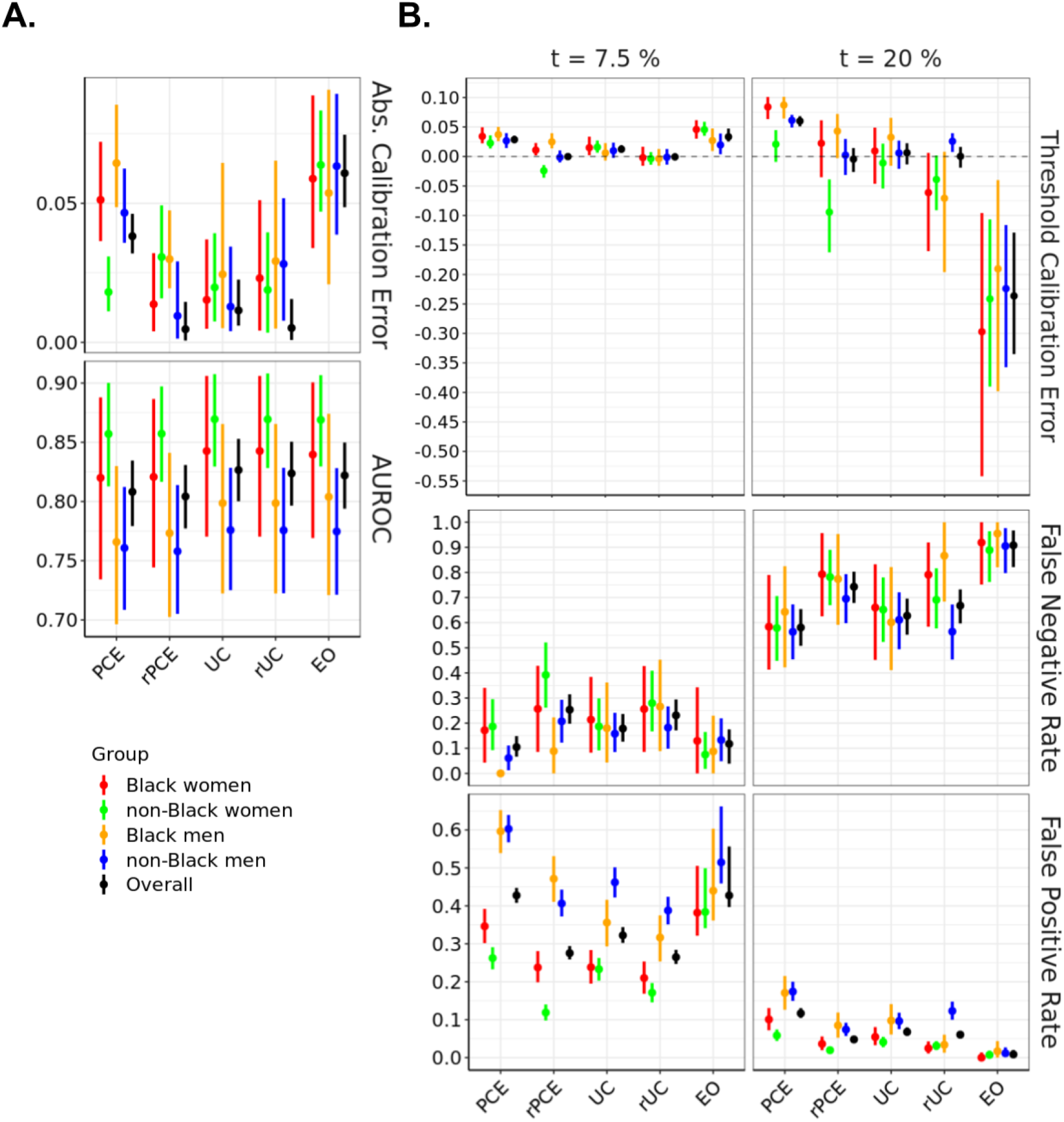
Model performance across evaluation metrics, stratified by demographic group, evaluated on the test set. PCE - original PCEs, rPCE - revised PCEs, UC - unconstrained model, rUC - recalibrated model, EO - Equalized Odds. The left panel shows AUROC and Absolute Calibration Error. The right panel shows False Negative Rates, False Positive Rates and Threshold Calibration Error at two therapeutic thresholds (7.5% and 20%).

**Figure 4.**
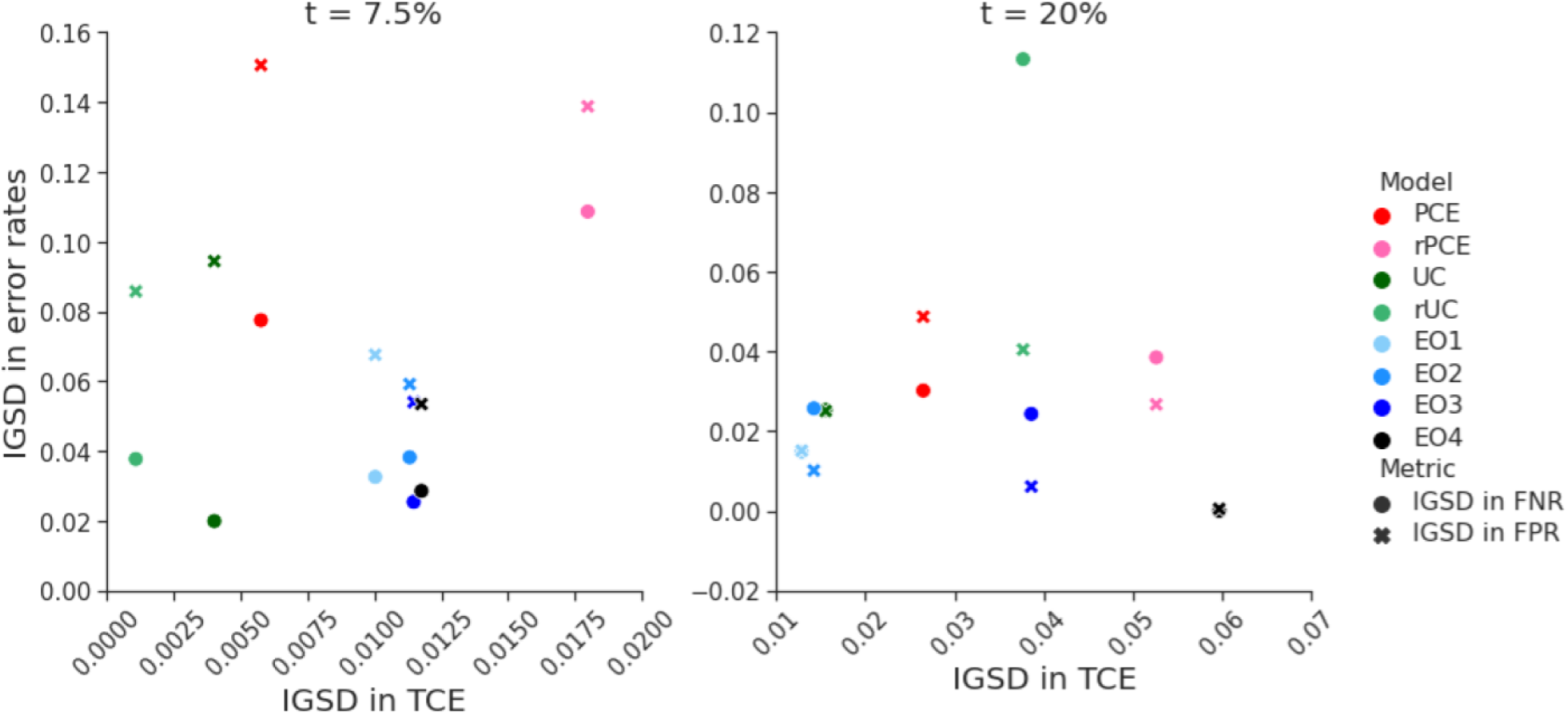
Relationship between inter-group variability in Threshold Calibration Rate (TCE) and error rates. The figure shows the relationship between inter-group standard deviation (IGSD) of threshold calibration error (on the x-axis) and IGSD of False Negative Rate (FNR, circles) and False Positive Rate (FPR, crosses) across the models: PCE - original PCEs, rPCE - revised PCEs, UC - unconstrained model, rUC - recalibrated model, EO1-4 - Equalized Odds with increasing values of λ. The EO3 corresponds to the EO model discussed in the Results section. In the models we trained, IGSD of TCE scales inversely with the IGSD of FNR and FPR.

### Study population

Overall, 25,619 individuals met the inclusion criteria, of whom 80% (N = 20,495) were assigned to the training set, and 10% (N = 2,562) to each the recalibration and test sets. Table 1 summarizes the mean age, ASCVD event incidence and frequency of censoring across the six datasets and four demographic groups. A cohort construction flowchart is included in Supplementary Figure B1.

**Table 1.** Cohort characteristics for patients who met inclusion criteria. Data are grouped by sex and race, as well as dataset. Each group of patients is described by four values: total number of individuals, mean age, censoring-adjusted incidence of ASCVD events within 10 years of the initial examination, and fraction of censored individuals. *ASCVD event incidence was calculated by weighing the number of positive- and negative-outcome uncensored individuals with the sum of their IPCW weights.

| study | N | age | ASCVD event incidence* | % censored | N | age | ASCVD event incidence* | % censored |
| --- | --- | --- | --- | --- | --- | --- | --- | --- |
|  | Black women |  |  |  | Black men |  |  |  |
| ARIC | 1,812 | 53.2 | 5.70% | 6.51% | 1,216 | 53.8 | 9.61% | 10.03% |
| CARDIA | 232 | 43.0 | 4.42% | 8.19% | 153 | 42.7 | 1.63% | 14.38% |
| CHS | 304 | 70.7 | 22.52% | 15.46% | 181 | 70.5 | 30.89% | 27.62% |
| JHS | 1,310 | 51.4 | 2.77% | 14.96% | 751 | 51.1 | 4.47% | 14.11% |
| MESA | 768 | 60.3 | 5.18% | 9.64% | 630 | 60.9 | 7.19% | 13.17% |
| all | 4,426 | 54.6 | 5.69% | 10.26% | 2,931 | 55.1 | 8.15% | 13.07% |
|  | non-Black women |  |  |  | non-Black men |  |  |  |
| ARIC | 4,815 | 53.9 | 2.54% | 3.30% | 4,383 | 54.5 | 7.17% | 4.86% |
| CARDIA | 289 | 42.7 | 0.39% | 6.23% | 333 | 42.5 | 0.90% | 6.91% |
| CHS | 1,848 | 70.7 | 20.18% | 15.58% | 1,169 | 71.0 | 32.00% | 17.45% |
| FHS OS | 828 | 46.4 | 2.61% | 1.81% | 856 | 47.1 | 8.67% | 3.86% |
| MESA | 1,913 | 60.5 | 3.81% | 7.68% | 1,828 | 60.8 | 6.67% | 10.07% |
| all | 9,693 | 57.4 | 5.95% | 6.47% | 8,569 | 56.9 | 10.36% | 7.67% |
|  | All |  |  |  |  |  |  |  |
| all | 25,619 | 56.5 | 7.54% | 8.28% |  |  |  |  |

### Model performance

The UC model achieved an overall AUROC of 0.827, (95% CI=[0.800, 0.853]), comparing favorably with PCE (0.808 [0.779, 0.835]) and rPCE (0.804 [0.777, 0.831]), while maintaining differences of AUROC between groups (Figure 3A). While UC had a slightly higher overall ACE (0.011 [0.006, 0.023]) than rPCE (0.005 [0.001, 0.015]), as well as a slightly higher local miscalibration at t_1_ (TCE(t_1_) 0.012 [0.006, 0.019] vs 0.000 [−0.004, 0.005]), IGSD(TCE, t_1_) and IGSD(TCE, t_2_) both reduced under UC (from 0.018 to 0.004, and 0.053 to 0.016, respectively) (Figure 4).

The group recalibration procedure (rUC) reduced the magnitude of TCE(t_1_) overall (−0.001 [−0.007, 0.006]), and for each group, relative to UC (0.012 [0.006, 0.019]) (Figure 3A). While recalibration improved TCE(t_2_) overall (0.00 [−0.019, 0.016] vs 0.006 [−0.013, 0.023]), it increased the magnitude of miscalibration of individual groups - for instance, shifting TCE(t_2_) from 0.033 [−0.016, 0.066] to −0.071 [−0.196, 0.008] for Black men, and increasing IGSD(TCE, t_2_) to 0.038. We also observe that, while TCE(t_1_) and IGSD(TCE, t_1_) improved for rUC, IGSD(FNR, t_1_) worsened, increasing from 0.020 to 0.038, as did IGSD(FPR, t_2_) and IGSD(FNR, t_2_) (Figure 4). Additionally, at each threshold, for all models, we observe a relationship between TCE, FPR and FNR: increased TCE (overestimation) leads to higher FPR and lower FNR, and decreased TCE (underestimation) - to lower FPR an higher FNR (Figure 3B).

The equalized odds procedure generated models with FPR and FNR which approached similar values across groups at both t_1_ or t_2_ - bringing IGSD(FPR, t_1_) to 0.054 from 0.094 (Figure 4) while maintaining almost identical AUROC to UC (0.822 [0.794, 0.8]) (Figure 3A). However, it did so by trading off error rates in opposite directions at the two thresholds, as described above (Figure 3B). It also increased the magnitude of TCE at both thresholds (from 0.012 [0.006, 0.019] to 0.033 [0.025, 0.047] at t_1_ and from 0.006 [−0.013, 0.023] to −0.236 [−0.335, −0.129] at t_2_), and increased IGSD(TCE, t_1_) to 0.011 (from 0.004) and IGSD(TCE, t_2_) to 0.039 (from 0.016), implying that the scores generated by the EO model did not closely correspond to their calibrated values.

## DISCUSSION

We identified local calibration of 10-year ASCVD risk prediction models at guideline-recommended thresholds as necessary for fair shared decision-making about statin treatment between patients and physicians. We find that the rPCEs [11] differ in local calibration between groups - making guideline-compatibility of rPCE inconsistent across groups. We note that global measurements of calibration, used previously to evaluate the PCEs [11,14], did not capture this difference, illustrating the importance of local calibration evaluation.

Recalibrating the model separately for each group increased compatibility with guidelines at low levels of risk, while increasing inter-group differences in error rates. Conversely, estimators learned with an equalized odds constraint would not be concordant with existing guidelines as a result of induced miscalibration. Thus, absent a contextual analysis, fairness approaches that focus on error rates can produce misleading results.

In our experiments, group-recalibration did not improve calibration at t=20%. This may be due to the small sample size of the recalibration set, as well as of individuals predicted to be at high risk. This suggests that group-recalibration may not always be desirable, especially if local calibration of the unconstrained model is deemed acceptable. However, improvement in local calibration observed at t=7.5% may be more relevant than calibration at higher risk levels for informing statin initiation decisions, since benefits of treatment are clearer at higher risk levels.

Several design choices may have impacted the results, including the use of a single model with race and sex as variables in the UC and EO models, the use of a logistic regression as a recalibration method, and the use of an in-processing method that focused on particular decision thresholds to impose equalized odds. We anticipate that alternative modeling choices would impact the size of the observed effects, but would likely not change the conclusions, since known statistical tradeoffs exist between equalized odds and calibration [18,21,22].

Given this analysis, we recommend that developers building models for use with the ACC/AHA guidelines prioritize calibration across a relevant range of thresholds, and report group-stratified evaluation of local calibration alongside metrics of global fit. Before a model is deployed in a new setting, we recommend that it be evaluated on the target population, stratified by relevant groups - and group-recalibrated, if necessary. Knowledge about local miscalibration should also be incorporated into risk calculators to actively inform the physician-patient shared decision making conversations, but should not replace recalibration efforts, since calibrated predictions are better suited for reasoning about potential consequences of treatment [10].

Our analysis inherits the assumptions about relative importance of relevant risks and benefits used to derive therapeutic thresholds (Supplement C), which often fail to consider the impact of social determinants of health on treatment efficacy and of structural forms of discrimination in generating health disparities [28]. Additionally, our use of self-identified racial categories - which can be understood as proxies for systemic and structural racist factors impacting health - may be inappropriate, potentially exacerbating historical racial biases and disparities in the clinical settings [29,30]. Derivation of new risk prediction models may be necessary for multiethnic populations [12]. Future work should explore decision analysis and modeling choices that incorporate this context.

## CONCLUSION

Our analysis is one of the first to consider algorithmic fairness in the context of clinical practice guidelines. It illustrates general principles that can be used to identify contextually relevant fairness evaluations of models used in clinical settings in the presence of clinical guidelines. Such analysis should include careful consideration of the interplay between model properties, model-guided treatment policy, as well as the potential harms and benefits of treatment, for each relevant subgroup. At the same time, we note that striving for model fairness is unlikely to be sufficient in addressing health inequities, especially when their sources lay upstream of the model-guided intervention, as is the case of structural racism [28]. We encourage future work to situate fairness analyses in this broader context.

## Data Availability

BioLINCC - links listed below.

https://biolincc.nhlbi.nih.gov/studies/cardia/

https://biolincc.nhlbi.nih.gov/studies/chs/

https://biolincc.nhlbi.nih.gov/studies/framoffspring/

https://biolincc.nhlbi.nih.gov/studies/jhs/

https://biolincc.nhlbi.nih.gov/studies/mesa/

https://biolincc.nhlbi.nih.gov/studies/aric/

## ACKNOWLEDGEMENTS

We thank Alison Callahan, Scotty Fleming, Julian Genkins, Nikolaos Ignatiadis, Jonathan Lu, Sherri Rose, Sanjana Srivastava, Ethan Steinberg, Jennifer Wilson, Yizhe Xu and Steve Yadlowsky for insightful comments and discussion. Approval for this non-human subjects research study is provided by the Stanford Institutional Review Board, protocol IRB-46829. This work is supported by the National Heart, Lung, and Blood Institute grant 5R01-HL144555 and the Stanford Medicine Program for AI in Healthcare. Any opinions, findings, and conclusions or recommendations expressed in this material are those of the authors and do not necessarily reflect the views of the funding bodies.

## AUTHOR CONTRIBUTIONS

AF and SRP conceived of the presented idea, and developed it with support from BP and NHS. AF cleaned and preprocessed the data, trained predictive models, generated and analyzed model evaluations. SRP provided code for training and evaluation and analyzed model evaluations. AF and SRP wrote the manuscript in consultation with BP and NHS. NHS supervised the project.

## Supplement A

### REGULARIZATION APPROACHES FOR EQUALIZED ODDS AT A THRESHOLD

Let 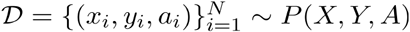 be a dataset, where *X* designates patient-level features, *Y* is a binary indicator for the occurrence of an outcome, and *A* s a discrete attribute that stratifies the population into *K* disjoint groups. We learn a model fθ(x) to estimate 𝔼 [*Y* | X = x ] = *P*(*Y* = 1 | *X* = *x*)using empirical risk minimization:

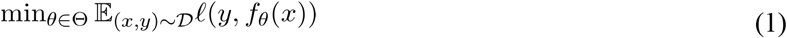

where ℓ is the cross-entropy loss and the expectation with respect to the dataset *𝒟* indicates an empirical average over the dataset.

To construct a training objective that penalizes violation of equalized odds, we add a regularizer *M*_EqOdd_ to the training objective, with the degree of regularization controlled by a non-negative scalar *λ*:

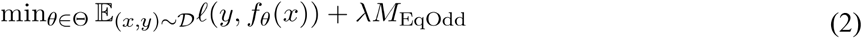

We derive a differentiable regularizer for this setting by first noting that the true positive rate at a threshold *t* can be defined as

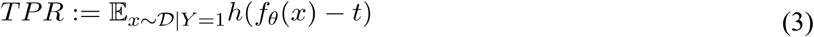

where 𝔼_(x,y) ∼ *𝒟* | *Y* = 1_ indicates the expectation over the subset of the population for which *Y* = 1, and *h(z)* = 1 {z > 0} is the step function. Similarly, the false positive rate is given by

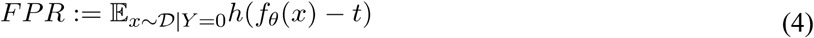

In order to achieve equal sensitivity (true positive rate) across groups and equal specificity (true negative rate) across groups, it is sufficient for the model to achieve a true positive rate and false positive rate for each group equal to the true positive rate and false positive rate of the population overall. The direct incorporation of the true positive rate and false positive rate, as formulated above, does not provide a useful signal for stochastic gradient descent when incorporated into a regularizer because the step function *h(z)* has a derivative of zero everywhere that the derivative is defined. To address this issue, we replace the step function *h(z)* with the sigmoid function 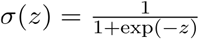, following Cotter et al. [1]. Furthermore, in practice, we compare the log of the scores to the log of the threshold. Terms representing the difference between the relaxed rates for a group *𝒜*_*k*_ with the population are given by:

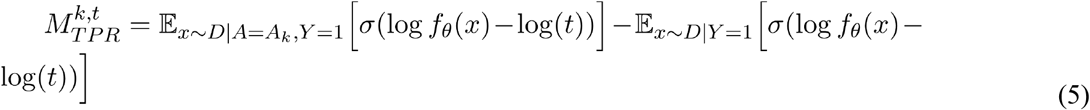

and

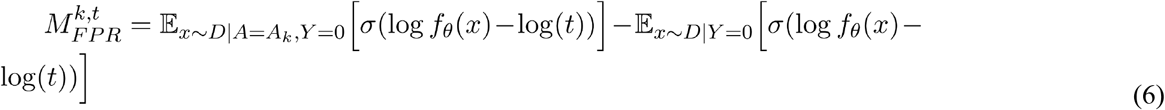

To construct the regularizer, we combine these terms into a single non-negative regularizer over all groups and thresholds:

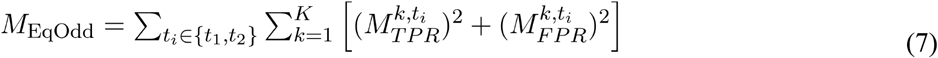

To adjust for censoring, we use an inverse probability of censoring (IPCW) approach, and replace the empirical averages in Equations 1, 2, 5, and 6 with weighted ones that incorporate the IPCW weights.

## Supplement B

**Supplementary Figure B1.**
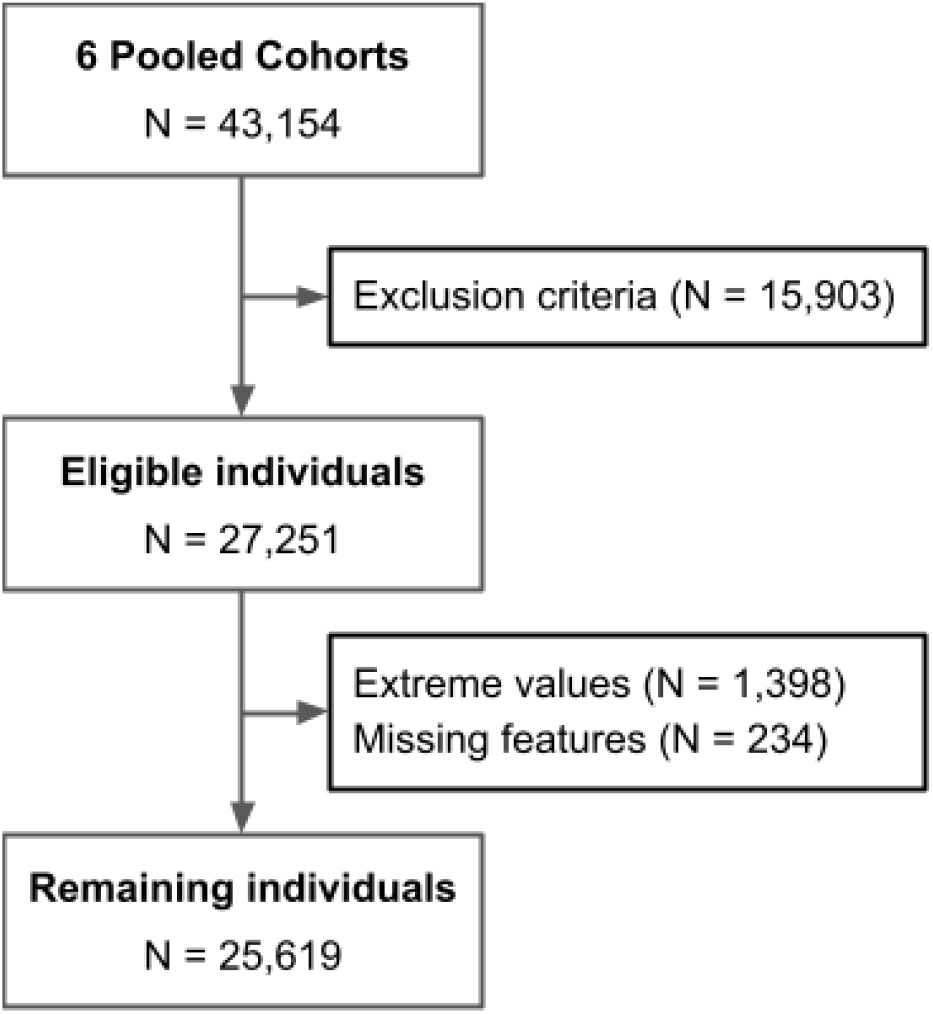
Cohort construction flowchart. We excluded individuals younger than 40 or older than 79, those with past history of myocardial infarction, stroke, coronary bypass surgery or angioplasty, congestive heart failure or atrial fibrillation, and those who were receiving statins at the time of the initial exam. We removed records with extreme values of systolic blood pressure (outside 90-200 mmHg), total cholesterol and high-density lipoprotein cholesterol (outside 130-320 and 20-100 mg/dL, respectively) and those with missing features.

**Supplementary Figure B2.**
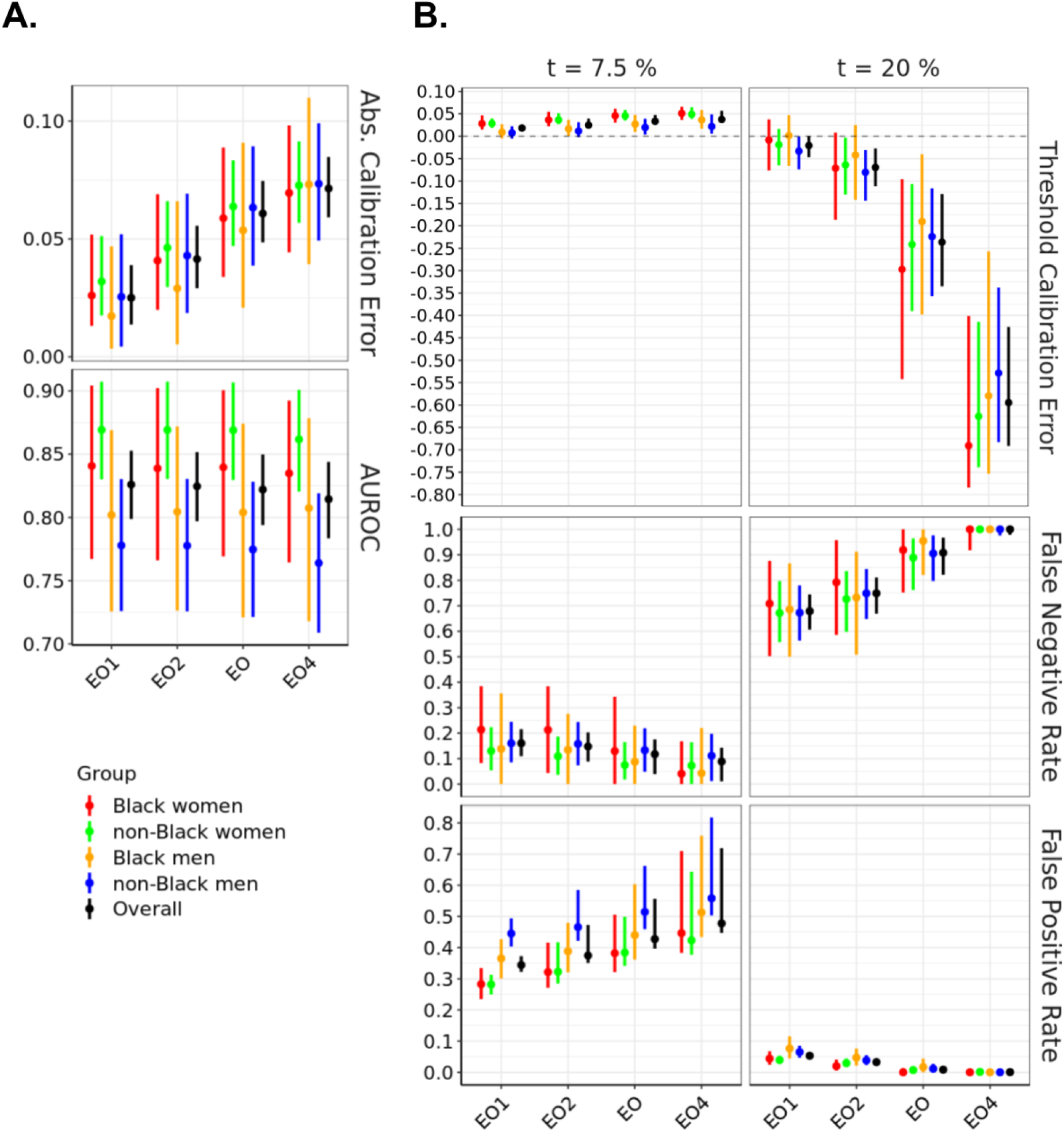
Performance of equalized odds models with different settings λ, stratified by demographic group, and evaluated on the held out test set. EO1 - Equalized Odds (λ=0.1), EO2 - Equalized Odds (λ=0.215), EO - Equalized Odds (λ=0.464), EO4 - Equalized Odds (λ=1). The left panel shows AUROC and Absolute Calibration Error. The middle panel shows Threshold Calibration Error at two therapeutic thresholds (7.5% and 20%). The right panel shows False Negative and False Positive Rates at the two therapeutic thresholds.

## Supplement C

### THE MEANING OF THRESHOLDS

The American College of Cardiology and the American Heart Association (ACC/AHA) 2019 guidelines on primary prevention of atherosclerotic cardiovascular disease (ASCVD) recommend different statin treatment regimens based on an estimate of ASCVD risk paired with therapeutic thresholds of 7.5% and 20% [1,2]. When grounding fairness analysis in threshold-based clinical guidelines, it is important to consider the assumptions underlying those thresholds, which can have implications for fairness.

The 7.5% treatment threshold for individuals without diabetes or clinical ASCVD and non-elevated LDL-C was established as a risk level above which net absolute benefit is positive. This was based on evidence from multiple RCTs, which studied the impact of statin treatment on ASCVD risk reduction [2] (Supplementary Section 7.3). The guidelines quantified net absolute benefit as a difference between benefits and harms, with *benefit* defined as as the number of patients who would need statin treatment to prevent one ASCVD event over 10 years (number needed to treat, NNT), and *harm* the number of statin-treated patients who would yield one excess case of diabetes over 10 years (number needed to treat to harm, NNH), both of which depend on 1) the estimate of a 10-year ASCVD risk of a patient, as well as 2) risk reduction from statin dose.

In 2013, The ACC/AHA committee [2] considered 5% and 7.5% as therapeutic thresholds. Those numbers corresponded to 10-year prevalence of ASCVD in the control groups of relevant primary prevention RCTs (Air Force/Texas Coronary Atherosclerosis Prevention Study [AFCAPS/TEXCAPS], Collaborative Atorvastatin Diabetes Study [CARDS], Justification for the Use of Statins in Primary Prevention: An Intervention Trial Evaluating Rosuvastatin [JUPITER], and Management of Elevated Cholesterol in the Primary Prevention Group of Adult Japanese [MEGA]). The NNT and NNH for different statin doses were then compared at the thresholds. At 7.5%, NNH was higher than NNT for both moderate and high-intensity statin treatment - suggesting that treating patients with either dose would prevent more ASCVD events in more people than it would induce new cases of diabetes in. This observation led the committee to select 7.5% as an acceptable therapeutic threshold.

This threshold is not derived from an economic sense of utility, where the relative value of harms and benefits are accounted for. While the guidelines explicitly state that the harm of an ASCVD event is more significant than that of a new diabetes diagnosis, the relative weight of NNH and NNT is not quantified. If NNH and NNT were treated as equivalent, the optimal decision threshold would correspond to a risk value where NNH=NNT - and it would have been lower than 7.5% (∼2.5% for moderate intensity statins, and ∼6% for high intensity statins). The committee made their final recommendation based on the interpretation of the best available data and methods at the time. Therefore, the 7.5% threshold could be considered reflective of expert consensus rather than an algorithmic optimization.

The 20% threshold was added in the 2018 Cholesterol Clinical Practice Guidelines (Section 4.4.2) [1] as a risk value above which high-intensity statin therapy should be considered (with 7.5-20% range considered more appropriate for considering moderate- or high-intensity therapy). Further analysis of trial outcomes conveyed a relationship between proportional reduction in LDL cholesterol from baseline and reduction in ASCVD. Thus, a higher threshold was meant to convey the importance of higher intensity therapy to provide maximal benefit for high risk patients. This was enacted by introducing an additional decision threshold in the guidelines. Additionally, the 2019 Guidelines [5] included LDL-C reduction goals, rather than statin doses, for the different risk categories.

While we have heard anecdotal evidence in our conversations with physicians which implied that the thresholds were set to account for miscalibration, the guidelines imply that thresholds were not set explicitly to account for known miscalibration of the PCE models.

The guidelines also make several other assumptions, listed below, which are detailed in Table 5 of the Supplement to 2013 ACC/AHA guideline on the treatment of blood cholesterol [2].

- Similar relative risk reduction (RRR) for CVD events across patient groups.
- RRR is proportional to the degree of LDL-C lowering by statin therapy.
- Absolute benefit in risk reduction is proportional to baseline risk of group/individual and the intensity of statin therapy - patients and groups with higher absolute risk will get more absolute benefit from statins.
- Absolute risk for adverse outcomes is proportional to baseline risk of a given group, and the intensity of statin therapy.
- Similar relative risk reduction (RRR) for CVD events across patient groups.
- RRR is proportional to the degree of LDL-C lowering by statin therapy.
- Absolute benefit in risk reduction is proportional to baseline risk of group/individual and the intensity of statin therapy - patients and groups with higher absolute risk will get more absolute benefit from statins.
- Absolute risk for adverse outcomes is proportional to baseline risk of a given group, and the intensity of statin therapy.

Since their publishing, some of those assumptions have been reevaluated [6].

